# Risk factors for increased COVID-19 case-fatality in the United States: A county-level analysis during the first wave

**DOI:** 10.1101/2021.02.24.21252135

**Authors:** Jess A. Millar, Hanh Dung N. Dao, Marianne E. Stefopulos, Camila G. Estevam, Katharine Fagan-Garcia, Diana H. Taft, Christopher Park, Amaal Alruwaily, Angel N. Desai, Maimuna S. Majumder

## Abstract

The ongoing COVID-19 pandemic is causing significant morbidity and mortality across the US. In this ecological study, we identified county-level variables associated with the COVID-19 case-fatality rate (CFR) using publicly available datasets and a negative binomial generalized linear model. Variables associated with decreased CFR included a greater number of hospitals per 10,000 people, banning religious gatherings, a higher percentage of people living in mobile homes, and a higher percentage of uninsured people. Variables associated with increased CFR included a higher percentage of the population over age 65, a higher percentage of Black or African Americans, a higher asthma prevalence, and a greater number of hospitals in a county. By identifying factors that are associated with COVID-19 CFR in US counties, we hope to help officials target public health interventions and healthcare resources to locations that are at increased risk of COVID-19 fatalities.

## 1 Introduction

The Severe Acute Respiratory Syndrome Coronavirus 2 (SARS-CoV-2) originated in Wuhan, China in November 2019 and has since spread to 210 countries worldwide.^1^ By June 12th, 2020, SARS-CoV-2 had caused over 2 million Coronavirus Disease 2019 (COVID-19) cases and 114,753 deaths in the United States (US).^2,3^ The distribution of infected cases and fatalities in the US has been heterogeneous across counties,^4^ and identification of sub-populations at risk of increased morbidity and mortality remains crucial to effective response efforts by federal, state, and local governments.^5^ Counties where governing officials are aware that their populations are at a higher risk of COVID-19 mortality, meaning the population experiences a higher case-fatality rate, may opt to tailor state policies or take earlier action to curtail the spread of SARS-CoV-2. Additionally, the federal government may opt to target vaccine resources to counties experiencing higher COVID-19 mortality rates.

The case-fatality rate (CFR) is defined as the number of deaths divided by the total number of confirmed cases from a given disease.^6^ When a disease is non-endemic, the CFR fluctuates over time. During the beginning of an epidemic, there is often a lag when counting the number of deaths compared to cases and hospitalizations, leading to an underestimation of the CFR. Furthermore, CFR will fluctuate rapidly early in an epidemic when each additional case or death has an excessive impact on calculating CFR. It is important to not only account for the lag between cases and deaths (i.e., lag-adjusted CFR), but also to ensure that the CFR is no longer fluctuating.

In this study, we use a lag-adjusted CFR to conduct a county-level mortality risk factor analysis of demographic, socioeconomic, and health-related variables in the US during the first wave of the COVID-19 pandemic (March 28, 2020 to June 12, 2020). We expand upon prior work by considering possible risk factors of an increased CFR from multiple categories (e.g., non-pharmaceutical interventions such as shelter-in-place orders,^7^ prevalence of pre-existing conditions such as cardiovascular disease,^8^ and socio-economic circumstances such as hospital accessibility^9^) in a single model.

## 2 Methods

All code for our work can be found on our GitHub repository.^10^

### Study Population

We conducted a cross-sectional ecological study to assess risk factors associated with an increased COVID-19 lag-adjusted CFR in US counties. Our study population included 3,004 counties or county-equivalents with Federal Information Processing Standards (FIPS), a unique code for US federal identification. Only publicly available aggregate data were used; therefore, no IRB approval was required.

### County-Level Variables

We identified potential risk factors across several different categories: demographic, socioeconomic, healthcare accessibility, comorbidity prevalence, and non-pharmaceutical interventions. Each category targeted variables relevant to the risk of COVID-19 mortality by conducting a comprehensive literature search on March 28, 2020 supplemented with variables relevant to other respiratory epidemics (Appendices A1-2).^11–14^ Only variables with publicly available data sources at the county-or state-level were included. Appendix A3^14^ lists data sources, variable descriptions, and manipulations (if applicable). We directly imported and cleaned the datasets using R (v3.6.3).

### Case-Fatality Rate (CFR) Data and Calculation

To calculate CFR during the first COVID-19 wave in the US, we obtained open access county-level COVID-19 data from the New York Times through June 12, 2020, the date the CDC released guidance for easing restrictions as states began to reopen.^2,15^ Only data that contained FIPS county codes to identify case and death locations were included. County-level data for New York City, NY was accessed from the New York City Department of Health and Mental Hygiene.^16^ To calculate lag-adjusted CFR (laCFR), we used Nishiura et al.’s method, expanded upon by Russell et al., to account for the delay between COVID-19 diagnoses and deaths.^17,18^ We updated this approach by using time-from-hospitalization-to-death from the US population.^18,19^ Details can be found in Appendix A4.^17,18^ The final dataset included 1,779 counties with 1,968,739 cases and 106,279 deaths, comprising 96.8% of national cases and 96.8% of national deaths as of June 12, 2020.

### Statistical Analysis

To reduce multicollinearity, we eliminated linear combinations and variables with correlations >0.5 using the R package caret (v6.0.86). Remaining variables were screened for missingness and missing values were imputed using five imputations in the R package mice (v3.8.0). Data were randomly split into training (1,186 counties) and testing sets (593 counties) to assess generalizability (a table of the characteristics can be seen in Appendix A5). A negative binomial linear model with an offset for the number of COVID19 cases per county was chosen based on Kolmogorov-Smirnov and dispersion tests found in the R package DHARMa (v0.3.1). Variable selection was conducted using purposeful selection, an iterative process in which covariates are removed from the model if they are neither significant nor confounders.^20^ With clinical risk factors, purposeful selection outperforms other variable selection procedures and tests for the presence of confounders.^20^ Removing highly correlated variables beforehand reduces the chance of multicollinearity between non-significant variables that may have been retained in purposeful selection due to confounding effects. Per Bursac et al., we used the 0.1 α-level for initial selection using bivariate models and a change of >20% in any remaining model coefficients compared with the full multivariate model for confounding evaluation.^20^ All variables in the final model were significant at the 0.05 α-level, and no statistical confounders were included in the final model.

## 3 Results

Of the 64 variables collected, 22 were retained for analysis after minimizing correlation (Appendix A3-7^14^). Multiple imputation was used to correct for missingness (less than 2%) in two of the retained variables, neither of which appeared in the final model.^21^ Fifteen variables were significant in bivariate models in the first step of purposeful selection, and were included in the initial multivariate model. Eight variables were significant in the initial multivariate model and were retained in the final model. Including variables that were non-significant in the bivariate models with these eight variables did not significantly change the performance of the model, as determined by the Likelihood Ratio Test. No potential confounders were identified among the correlation minimized variables that were previously discarded due to non-significance in the models.

The final model is shown in Exhibit 1. The negative binomial model appears to be a good fit, capturing the mean-variance relationship observed in the data and displaying expected residuals (Exhibit 2A and 2B). The model was well-calibrated, with the training and testing model having comparable coverage and relative Gini score (Exhibit 2C and 2D; Appendix A7). Since we used a negative binomial model with an offset, the exponentiated coefficients represent the change in laCFR observed for a one-unit increase in each continuous variable, assuming all other variables in the model are held constant (Exhibit 3). Four variables were inversely associated with laCFR: number of hospitals per 10,000 people (−39% laCFR per additional hospital per 10,000), banning religious gatherings during the initial state or county shutdown (−13% laCFR if religious gatherings were banned), percentage of housing units that were mobile homes (−0.79% laCFR per 1% increase in the proportion of mobile homes), and percentage of population without health insurance (−1.5% laCFR per 1% increase in percentage uninsured). Four variables were directly associated with laCFR: percentage over age 65 (+4.4% laCFR per 1% increase in population over age 65), percentage Black or African American (BAA) (+0.97% laCFR per 1% increase in BAA population), percentage with asthma (+9.1% laCFR per 1% increase in asthma prevalence), and number of hospitals (+3.1% laCFR per one additional hospital). Exhibit 4 demonstrates the relationship between each variable and the laCFR over a range of values.

**Exhibit 1.**
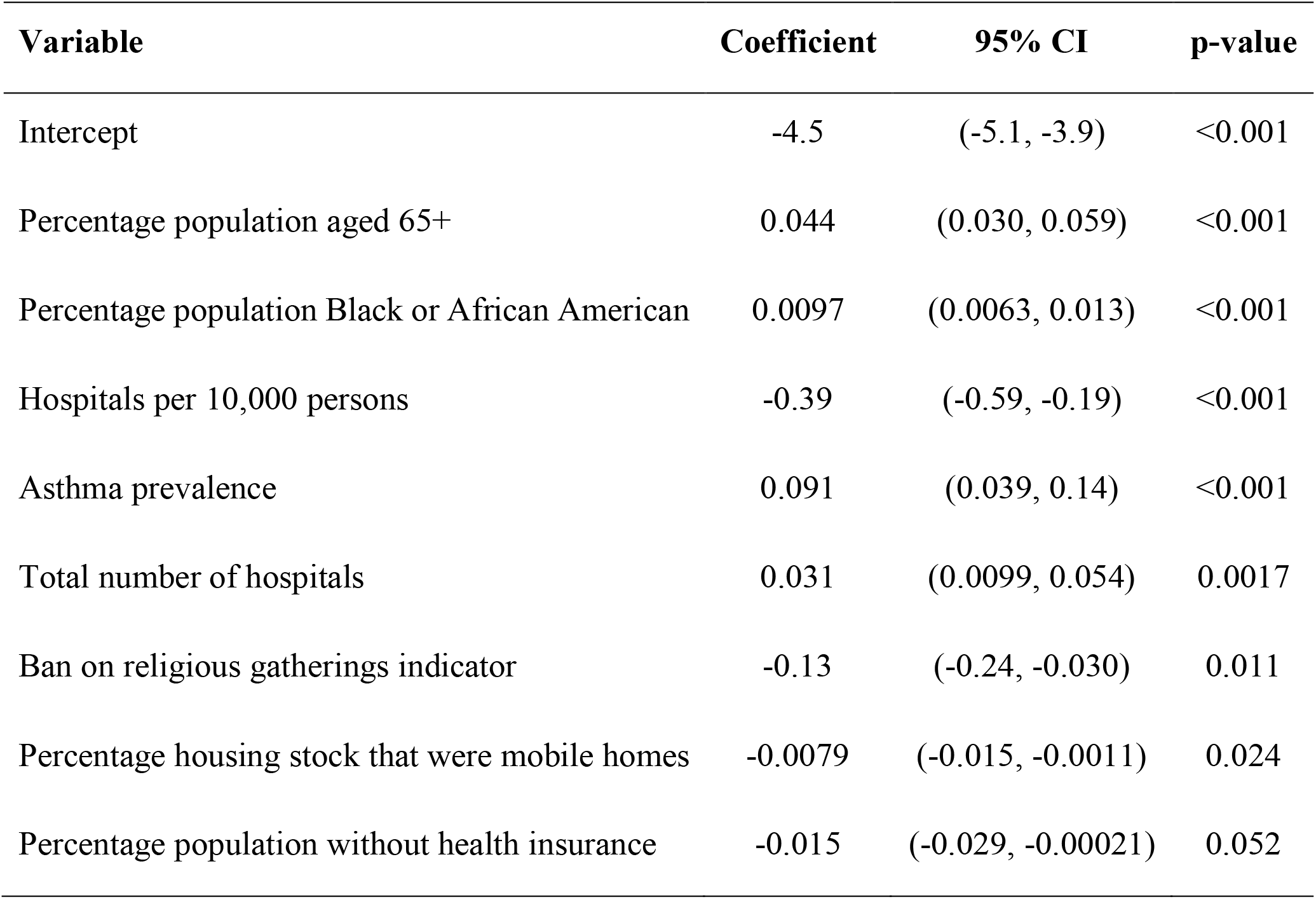
Parameter estimates for the final multivariate model of laCFR.

**Exhibit 2.**
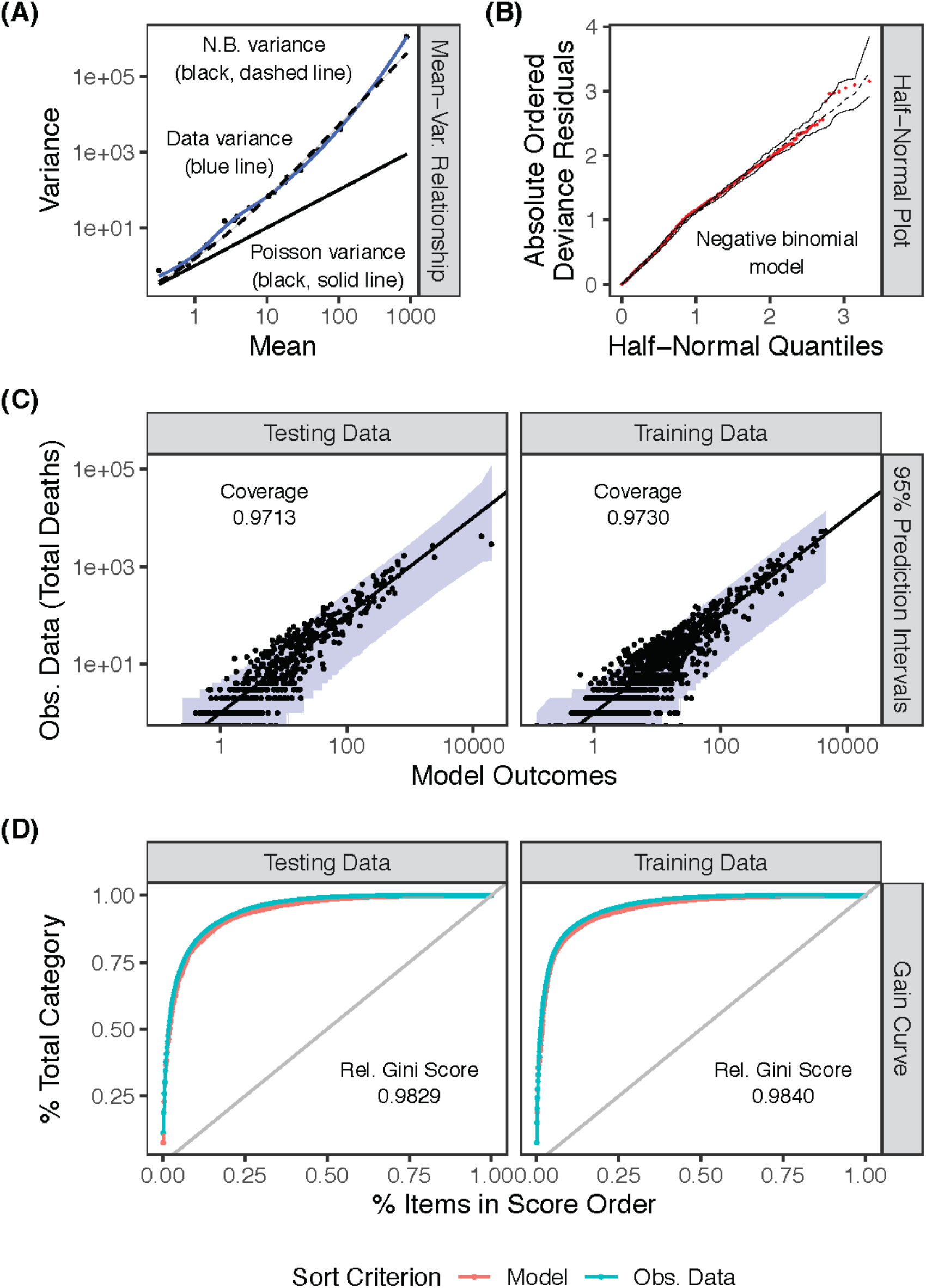
Assessing model fit. Plots showing (A) mean-variance relationship of the observed county-level COVID-19 laCFRs, (B) half-normal residuals, (C) model outcomes found within the prediction intervals for training data and testing data for the county-level COVID-19 laCFRs, and (D) gain curves for training data and testing data for the county-level COVID-19 laCFRs.

**Exhibit 3.**
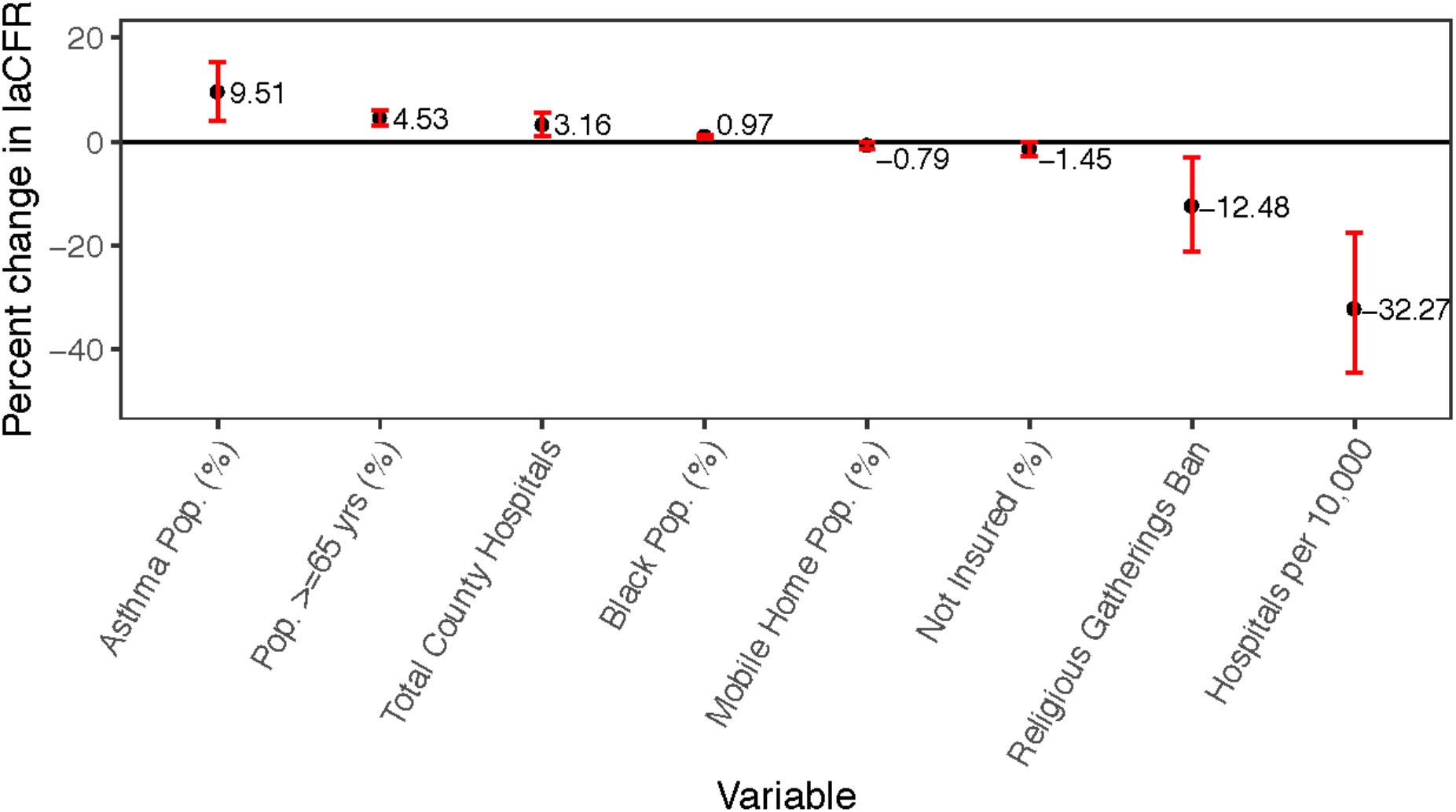
Percentage change in COVID-19 laCFR given a 1 unit increase in the variable for each individual variable (shown in black dots) and 95% confidence interval (shown in red), using training data.

**Exhibit 4.**
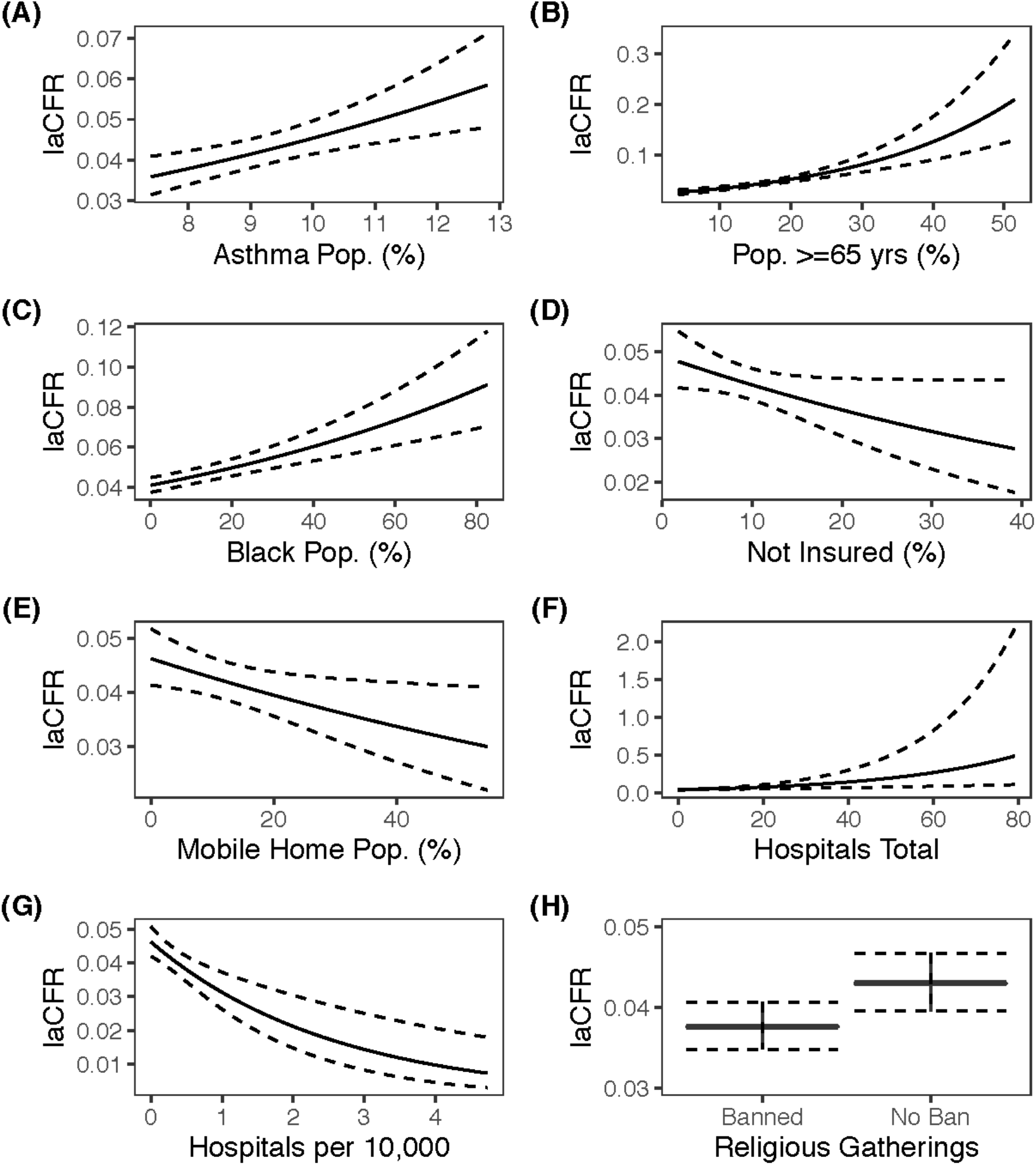
Relationship of each individual significant variable with COVID-19 laCFR over a range of values. To obtain estimates, all other variables were set to their median value in the training data, and the banned religious gatherings variable was set to 0 (indicating religious gatherings were not banned in a county, which account for half of counties included, Appendix A1). Means are shown as solid line black, and 95% prediction intervals are shown as dotted black lines.

## 4 Discussion

In this ecological study of mortality due to SARS-CoV-2 infection during the first wave of COVID-19 in the US, we found that county-level laCFR was significantly associated with eight variables. Four variables – banning religious gathering, proportion of mobile homes, hospitals per 10,000 persons, and proportion of uninsured individuals in a county – were associated with decreased laCFR. Four variables – percentage of population over age 65, total number of hospitals per county, prevalence of asthma, and percentage of population BAA – were associated with increased laCFR. Each variable provided unique insights into factors that may be worth considering for county-level COVID-19 response efforts.

### Inverse Association with Case-Fatality Rate

Our model indicated a 13% reduction in the average laCFR for counties that banned religious gatherings compared to counties that did not. Gatherings often involve dense mixing of people in a confined space, sometimes over long periods of time,^22^ which drives COVID-19 transmission.^23^ Interventions targeting increased physical distancing and limiting contact were introduced in some countries, including the closure of schools, places of worship, malls, and offices.^22^ Our model suggests that specifically exempting religious gatherings from bans may increase the laCFR, consistent with the combination of findings that (1) religious gatherings across the globe were linked to COVID-19 superspreader events^24^ and (2) older Americans (who are more likely to attend religious services than younger Americans^25^) are at increased risk of death due to COVID-19.

The percentage of the population living in mobile homes was also associated with a decrease in laCFR. A 1% increase in mobile home living was associated with a 0.79% decrease in laCFR. While a small difference at first glance, it becomes more meaningful when considering the large variation in mobile home living across counties. Between counties at the 25th percentile of percentage living in mobile homes (4%) and 75th percentile (18%), the difference in the percentage of mobile home living correlated with an 11% decrease in laCFR. This might represent a built-environment effect, given that mobile homes have separate plumbing and ventilation unlike apartments and other multi-family residences. Recent work suggests that fecal aerosol transmission of SARS-CoV2 can occur.^26^ Ventilation patterns in apartment complexes represent additional opportunities for transmission.^27^ The benefit of separate units such as mobile homes may be especially important to low-income workers who are both more at risk of death from COVID19 due to increased chance of having a co-morbid condition and more likely to live in multi-family housing with maintenance issues.^28^

The number of hospitals per 10,000 was also inversely associated with laCFR. We found that for each additional hospital per 10,000 inhabitants, the laCFR decreased by 39%, despite the exclusion of other healthcare-related variables due to non-significance (e.g., ICU bed availability). Prior work demonstrated that the percentage of ICU and non-ICU beds occupied by COVID-19 patients directly correlated with COVID-19 deaths,^29^ and a county with more hospitals per 10,000 inhabitants may be able to cope with more COVID-19 cases before reaching the same percentage of hospitals beds occupied as a county with fewer hospitals per 10,000 inhabitants. Furthermore, because adding beds requires fewer resources than adding hospitals, the number of hospitals per 10,000 persons in a county might represent a greater ability to expand capacity. As a result, using hospitals per 10,000 may be a better indicator of healthcare capacity than the number of ICU beds early on in the pandemic. Because healthcare resources in the US correlate with community wealth,^30^ the rate of hospitals per 10,000 may also reflect increased community wealth and the protective effect of higher socio-economic status on health. More hospitals per 10,000 persons may also represent increased competition for patients, which is associated with decreased mortality from community-acquired pneumonia.^31^

Unexpectedly, the percentage uninsured was inversely associated with laCFR. We found a 1.5% reduction in laCFR for every 1% increase in uninsured inhabitants. Prior studies found longer travel times to COVID-19 testing facilities were directly associated with percentage uninsured.^32,33^ Because uninsured persons may be unable to readily access testing, this finding may relate to incomplete reporting, such that only individuals who survive long enough are tested for COVID-19, leading to a potential undercount of deaths attributable to SARS-CoV-2 infection.

### Direct Association with Case-Fatality Rate

In our model, a 1% increase in the population over 65 years old was associated with a 4.4% increase in average laCFR. This is consistent with recent epidemiological studies demonstrating an association between the severity of COVID-19 infection and age. According to provisional death data from the National Center of Health Statistics, people aged 65 and older have a 90-to 630-fold higher risk of mortality due to COVID-19 than 18-29-year olds.^34^

Also directly associated with laCFR was the total number of hospitals per county, with an observed increase of 3.1% in average laCFR per additional hospital. This variable was strongly correlated with total population (r=0.92). Given that the number of hospitals per 10,000 was associated with decreased laCFR, this correlation suggests that total hospitals might be a proxy indicator for total population. Previous work assessed population density as a risk factor for increased laCFR, but not total population.^23^ Since our analysis focused on the first wave of COVID-19, this variable could reflect overwhelmed healthcare systems in highly populated counties where most of the COVID-19 cases initially occurred.^35^

Asthma prevalence was also directly associated with laCFR. A 1% increase in asthma prevalence was associated with a 9.1% increase in laCFR. Evidence regarding asthma as a risk factor in COVID-19 is mixed. Although the US CDC has determined that patients with moderate to severe asthma belong to a high-risk group,^36^ the Chinese CDC indicated that asthma was not a risk factor for severe COVID-19.^37^ One study showed that COVID-19 patients with asthma were of older age and had an increased prevalence of multiple comorbidities compared to those without asthma,^38^ but that the presence of asthma alone was not a risk factor for increased mortality.^38^ Thus, despite our findings, it is unclear whether asthma has a direct impact on COVID-19 disease or if other factors may be associated with both asthma and COVID-19. One such potential confounder is exposure to air pollution, as air pollution is associated with both asthma and risk of death from COVID-19.^39^

Finally, laCFR was directly associated with the percentage of the population identifying as BAA in a county. Our model showed that a 1% increase in BAA was associated with a 0.97% increase in the laCFR. This likely reflects the effects of structural racism in the US, where BAAs have fewer economic and educational opportunities than White Americans and as a result are exposed to increased risk of morbidity and mortality from COVID-19.^40^ Dalsania et al. also found that the social determinants of health contributed to a unequal impact of the COVID-19 pandemic for BAA at the county level.^41^ A study by Golestaneh showed that US counties with BAA as the majority had three times the rate of infection and almost six times the rate of death as majority White counties.^42^ Factors underlying this trend include years of structural racism resulting in a lack of financial resources, increased reliance on public transportation, housing instability, and dependence on low-paying retail jobs.^43^ Our approach considered several other variables that might explain the effect but were either non-significant (e.g., household crowding, percentage of households without a vehicle, and county land area) or were correlated with percentage BAA (e.g. percentage single parent households and percentage living in poverty), further emphasizing the role of systemic racism in COVID-19 laCFR.

### Excluded Predictor Variables

In reducing multicollinearity and using purposeful selection, several variables were surprisingly excluded. One of these excluded variables was population density, although higher population density had been hypothesized to increase contact rate and non-adherence with physical distancing.^23^ Diabetes and cardiovascular disease were excluded, despite multiple studies reporting these conditions as risk factors for COVID-19 mortality.^44,45^ While these factors are important at an individual level to assess the mortality risk, our model suggests that other variables may be more informative at the county-level, underscoring the value of ecological studies.

### Study Strengths and Limitations

This study had several strengths besides the benefits of an ecological design when considering population interventions. First, the data were nationally representative, including over 50% of all US counties. Our model captured the variability in the data and accounted for the observed data distribution. The model also captured almost all outcomes within the prediction interval for both training and testing data sets, with similar accuracy between them, which indicates that our model is generalizable within the US (Appendix A7). Additionally, our model based laCFR calculations on the distribution of times from hospitalization to death from US data,^19^ which differed from earlier Chinese data.^37^ Using US-based distribution of times likely improved our laCFR estimation for this study. The final model included several variables previously attributed to higher laCFR (such as older age)^34^ and included a variable unique to the pandemic shutdown, i.e., banning religious gatherings, giving more nuanced insights into heterogeneous COVID-19 mortality rates across counties.

Despite these strengths, our study had several limitations. First, under-reporting of cases might affect the accuracy of CFR calculation.^46^ The reported cases and deaths we used likely underestimated the true COVID-19 parameters. This underestimation was more among the asymptomatic and mild cases due to limited testing capacity and changes in testing practice; hence, the laCFR might have appeared inflated. Second, the type and timing of the tests used may have impacted the measured laCFR. Samples collected early during the infection can yield higher false negatives with RT-PCR tests.^47^ False negatives in critically-ill patients who later die could decrease the measured laCFR unless probable COVID-19 deaths are reported, while false negatives in mild cases who are not retested later could increase the measured laCFR as survivable cases go undetected. These are challenges for any CFR study and highlight the ongoing need for improved COVID-19 testing. Third, COVID-19 reporting practices vary widely by state. For example, Florida was found to report fewer COVID-19 deaths in the official tally than the Medical Examiners Commission.^48^ In addition to deliberate underreporting of deaths, states also vary in reporting of probable cases and deaths.^49^ Without national standards in the COVID-19 response, comparing case counts and deaths across state line–let alone county–is deterred by lack of clarity about how these data differ.^49^

Beyond these, our study was also limited by the fact that relevant data were frequently unavailable, including data on non-pharmaceutical interventions (NPI) and comorbidities. To limit missingness in the NPI data, we used state-level data when available given that counties also enforce state-level orders. However, there may be heterogeneity between county- and state-level information making this a less effective approach. Other variables of interest were not available at the state- or county-level, including information on contact tracing efforts and community compliance with public health mandates. Funding to collect public health information on more variables at a granular level would improve the information available to guide decision-making during emergencies. Another limitation was the highly correlated nature of the 64 variables considered for inclusion. Multicollinearity greatly affects the interpretability of coefficients and is rarely accounted for in epidemiologic studies.^50^ Highly correlated variables in a model are unstable and can bias standard errors, leading to unreliable p-values and unrealistic interpretations.^50^ Because we ensured our model interpretability by excluding highly correlated variables, not all of our collected 64 variables were screened for inclusion in the final model.

Finally, our study period ended in mid-June. This date was chosen because (1) enough cases had occurred in the US to obtain reliable estimates of laCFR by county and (2) it preceded CDC reopening guidance and a shift in reporting to the HHS Protect system, which is less readily available to the public than the prior CDC reporting system.51 The decision by the government to switch to the HHS Protect system hinders the ability of academic scientists to aid in the response to the on-going pandemic.^51^ Making these data more readily available to the public would permit inclusion of additional data for future research.

## 5 Conclusion

This study highlights several variables that were associated with county-level laCFR during the first wave of COVID-19 in the US. Though further research is needed to examine the effects of additional NPIs, our work provides insights that may aid in targeting response and vaccination efforts for improved outcomes in subsequent waves.

## Data Availability

All data used was taken from publicly available data sets. The list of these data sets can be found in Appendix A3.

## 7 Author Contributions

JAM, HDND, DHT, AND, and MSM designed the study. JAM and HDND conducted data analysis. JAM, HDND, MES, CGM, KFG, DHT, CP, and AA cleaned the data and drafted the initial manuscript. All authors edited, read, and approved the final manuscript.

## 9 Acknowledgments

This project is part of the COVID-19 Dispersed Volunteer Research Network (COVID-19-DVRN) led by M. Majumder and A. Desai. The authors thank Marie Charpignon, Catherine Pollack, and Emily Ricotta for their thoughtful feedback and review of the manuscript.

