## Appendices A1 A7 for "Risk factors for increased COVID-19 case-fatality in the United States: A county-level analysis during the first wave"

### Appendix

PAGE

**Appendix Exhibit A1: County-level variables methods**

We included five demographic variables: total population, population density, the percentage of the population over age 65, the percentage of population 17 or younger, and race/ethnicity. All demographic variable data were from the 2018 American Community Survey 5-Year Data from the US Census annual survey,[1] except for race/ethnicity data from the U.S. Census Populations with Bridged Race Categories.[2]

We included 13 socioeconomic variables, with their data primarily from the 2018 American Community Survey 5-Year Data.[1] In addition to the commonly used socioeconomic variables, we included certain variables contributing to the composite Social Vulnerability Index (SVI). The SVI was created by the Centers for Disease Control and Prevention (CDC) to describe US geographic areas by their social vulnerability and has been validated by multiple studies within and outside of the CDC.[3-8] Social vulnerability is defined as "the characteristics of a person or community that affect their capacity to anticipate, confront, repair, and recover from the effects of a disaster."[4] We included individual SVI variables on socioeconomic status, household composition and disability, minority status and language, and housing and transportation. We preferred to use the individual variables rather than overall SVI or by theme because we were most interested in understanding which components of social vulnerability contributed to increased CFR.

We included 5 healthcare-related variables: number of hospitals per capita, number of ICU beds per capita, number of primary care physicians per capita, percentage of residents without health insurance, and percentage of Medicaid eligible residents. Variable data was from the Kaiser Health News,[9] the Heart Disease and Stroke Atlas,[10] and the 2018 American Community Survey 5-Year Data.[1]

We included 18 comorbidity variables: diagnosed diabetes prevalence; diagnosed obesity prevalence; hypertension hospitalization and death prevalence, cardiovascular disease (CVD), chronic obstructive pulmonary disease (COPD), asthma, and cancer; Medicare beneficiaries with heart disease percentage, current smokers prevalence, and stroke-related hospitalization and mortality prevalence. Variable data was from the US Diabetes Surveillance System,[11] the Heart Disease and Stroke Atlas,[10] the Behavioral Risk Factor Surveillance System,[12] and the State Cancer Profiles by the National Cancer Institute.[13]

Non-pharmaceutical intervention data (including information on closing of public venues such as restaurants, gathering size limits, complete lockdown of non-essential activity in the county, if religious gatherings were included in gathering size limits, shelter-in-place orders, and social distancing mandates) were extracted from the COVID-19-intervention GitHub page, an open source data-sharing platform and compiled by Keystone Strategy.[14] However, this resource does not cover all counties, thus missing data was supplemented from a variety of governmental executive orders and news articles detailed in Appendix 5. Variables with dates were transformed to how many days the event occurred after the first case in a county. States where an intervention never occurred were given a zero. Since

47% of all counties did not ban religious gatherings, data on when religious gatherings were banned in a county was transformed into an indicator variable (1 if the ban occurred, 0 if not).

**Appendix Exhibit A2: Justifications for variable inclusion**

| Variable Topic | Variable Code | Variable Description | Justifications |
|---|---|---|---|
| Comorbidities | como_allheartdis_hosp | Heart Disease Hospitalization Rate per 1,000 Medicare Beneficiaries, 65+ | This variable is associated with the severity of and mortality due to COVID-19.[15] |
| Comorbidities | como_allheartdis_mort | Heart Disease Death Rate per 100,000, 35+ | This variable is associated with the severity of and mortality due to COVID-19.[15] |
| Comorbidities | como_asthma | Age-adjusted prevalence of adults who have been told they currently have asthma | This variable is associated with the severity of and mortality due to COVID-19.[16] |
| Comorbidities | como_cancer5yr | Age adjusted incidence of all cancer-5-year prevalence | This variable is associated with the severity of and mortality due to COVID-19.[17] |
| Comorbidities | como_COPD | Age-adjusted prevalence of adults diagnosed with chronic obstructive pulmonary disease | This variable is associated with the severity of and mortality due to COVID-19.[18] |
| Comorbidities | como_cvd_hosp | Total Cardiovascular Disease Hospitalization Rate per 1,000 Medicare Beneficiaries, 65+ | This variable is associated with the severity of and mortality due to COVID-19.[15] |
| Comorbidities | como_cvd_mort | Total Cardiovascular Disease Death Rate per 100,000, All Ages | This variable is associated with the severity of and mortality due to COVID-19.[15] |
| Comorbidities | como_pdiabetes | Age-adjusted prevalence of adults aged 20+ years with diagnosed diabetes (in %) by county | This variable is associated with the severity of and mortality due to COVID-19.[19] |
| Comorbidities | como_htn_hosp | Hypertension Hospitalization Rate per | This variable is associated with the severity of and mortality due to COVID-19.[20] |

| | | | |
|---|---|---|---|
| | | 1,000 Medicare Beneficiaries, 65+ | |
| Comorbidities | como_htn_mort | Hypertension Death Rate per 100,000 (any mention), 35+ | This variable is associated with the severity of and mortality due to COVID-19.[20] |
| Comorbidities | como_ medicareheartdizprev | Prevalence (in %) of heart disease among Medicare beneficiaries | This variable is associated with the severity of and mortality due to COVID-19.[15] |
| Comorbidities | como_pobesity | Age-adjusted prevalence of adults aged 20+ years with obesity (in %) by county | This variable is associated with the severity of and mortality due to COVID-19.[21] |
| Comorbidities | como_smoking | Age-adjusted prevalence of adults who are current smokers (variable calculated from one or more BRFSS questions) | This variable is associated with the severity of and mortality due to COVID-19.[18] |
| Comorbidities | como_stroke_hosp | Stroke Hospitalization Rate per 1,000 Medicare Beneficiaries, 65+ | This variable is associated with the severity of COVID-19.[22] |
| Comorbidities | como_stroke_mort | Stroke Death Rate per 1,000, 35+ | This variable is associated with the severity of COVID-19.[22] |
| Demographics | demo_landarea | County land area in square meters | Historically, more rural areas saw a lower burden of infectious disease because smaller populations meant diseases were less likely to be circulating,[23] suggesting that counties with smaller populations or larger land areas may be less impacted if those who are unwell come into contact with fewer people allowing the disease to burn out before cases and CFR climb. |
| Demographics | demo_p60more | Percentage of population aged 60 years or older | COVID-19 has a higher fatality rate for older populations, while sparing younger ages from more severe forms of the disease.[24] |

| | | | |
|---|---|---|---|
| Demographics | demo_p65more | Percentage of population aged 65 years or older | COVID-19 has a higher fatality rate for older populations, while sparing younger ages from more severe forms of the disease.[24] |
| Demographics | demo_p45_64 | Percentage of population aged 45 to 64 | COVID-19 has a higher fatality rate for older populations, while sparing younger ages from more severe forms of the disease.[24] |
| Demographics | demo_popdensity | Population density | Population density may make social distancing more challenging and may also result in a higher effective contact rate.[25] |
| Demographics | demo_population | Total population of each county (same as demo_bridgedrace_total) | Historically, more rural areas saw a lower burden of infectious disease because smaller populations meant diseases were less likely to be circulating,[23] suggesting that counties with smaller populations or larger land areas may be less impacted if those who are unwell come into contact with fewer people allowing the disease to burn out before cases and CFR climb |
| Healthcare access & capacity | hc_hospitals | Number of Hospitals | Counties with greater healthcare resources available will presumably be able to manage a higher case-load before becoming overwhelmed.[26] |
| Healthcare access & capacity | hc_hospitals_per10000 | Number of Hospitals per 10000 | Counties with greater healthcare resources available will presumably be able to manage a higher case-load before becoming overwhelmed.[26] |
| Healthcare access & capacity | hc_icubeds_per1000 | Number of ICU beds per 1000 | Counties with greater healthcare resources available will presumably be able to manage a higher case-load before becoming overwhelmed.[26] |
| Healthcare access & capacity | hc_icubeds | Number of ICU beds per 1000 | Counties with greater healthcare resources available will presumably be able to manage a higher case-load before becoming overwhelmed.[26] |
| Healthcare access & capacity | hc_icubeds_per60more | Number of ICU beds per >60 year resident | Counties with greater healthcare resources available will presumably be able to manage a higher case-load before becoming overwhelmed.[26] |

| | | | |
|---|---|---|---|
| Healthcare access & capacity | hc_icubeds_per65more | Number of ICU beds per >65 year resident | Counties with greater healthcare resources available will presumably be able to manage a higher case-load before becoming overwhelmed.[26] |
| Healthcare access & capacity | hc_medicaid | Medicaid eligible | Uninsured Americans are less likely to access health care when needed, more likely to delay treatment, are at higher risk of hospitalization, and also more likely to have preventable illnesses or uncontrolled chronic illnesses, which may put them at higher risk of serious COVID-19 illness.[27,28] |
| Healthcare access & capacity | hc_Pnotinsured_acs | Percentage without Health Insurance | Uninsured Americans are less likely to access health care when needed, more likely to delay treatment, are at higher risk of hospitalization, and also more likely to have preventable illnesses or uncontrolled chronic illnesses, which may put them at higher risk of serious COVID-19 illness.[27,28] |
| Healthcare access & capacity | hc_primarycare | Number of primary care physicians in the county | Counties with greater healthcare resources available will presumably be able to manage a higher case-load before becoming overwhelmed.[26] |
| Healthcare access & capacity | hc_primarycare_per1000 | Primary Care Physician per capita | Counties with greater healthcare resources available will presumably be able to manage a higher case-load before becoming overwhelmed.[26] |
| Non-pharmaceutical intervention | npi_keystone_closing_of_public_venues | Government order closing venues such as restaurants, theaters, and bars | We expect non-pharmaceutical intervention (NPI) can have an influence on COVID-19 mortality. At the time of variable inclusion, there were no published results for COVID-19 NPI yet. |
| Non-pharmaceutical intervention | npi_keystone_gathering_size_10_0 | Gathering size limited to 10 or fewer people | We expect non-pharmaceutical intervention (NPI) can have an influence on COVID-19 mortality. At the time of variable inclusion, there were no published results for COVID-19 NPI yet. |
| Non-pharmaceutical intervention | npi_keystone_gathering_size_100_to_26 | Gathering size limited to 26 to 100 people | We expect non-pharmaceutical intervention (NPI) can have an influence on COVID-19 mortality. At the time of variable inclusion, there were no published results for COVID-19 NPI yet. |

| | | | |
|---|---|---|---|
| Non-pharmaceutical intervention | npi_keystone_gathering_size_25_to_11 | Gathering size limited to 11 to 25 people | We expect non-pharmaceutical intervention (NPI) can have an influence on COVID-19 mortality. At the time of variable inclusion, there were no published results for COVID-19 NPI yet. |
| Non-pharmaceutical intervention | npi_keystone_gathering_size_500_to_101 | Gather size limited to 101 to 500 people | We expect non-pharmaceutical intervention (NPI) can have an influence on COVID-19 mortality. At the time of variable inclusion, there were no published results for COVID-19 NPI yet. |
| Non-pharmaceutical intervention | npi_keystone_lockdown | Lockdown | We expect non-pharmaceutical intervention (NPI) can have an influence on COVID-19 mortality. At the time of variable inclusion, there were no published results for COVID-19 NPI yet. |
| Non-pharmaceutical intervention | npi_keystone_non_essential_services_closure | Government order closing non-essential services and shops | We expect non-pharmaceutical intervention (NPI) can have an influence on COVID-19 mortality. At the time of variable inclusion, there were no published results for COVID-19 NPI yet. |
| Non-pharmaceutical intervention | npi_keystone_Other | Other, unspecified, NPI | We expect non-pharmaceutical intervention (NPI) can have an influence on COVID-19 mortality. At the time of variable inclusion, there were no published results for COVID-19 NPI yet. |
| Non-pharmaceutical intervention | npi_keystone_religious_gatherings_banned | Cancellation of religious gatherings either explicitly or implicitly through gathering size limitations that do not exempt religious services | We expect non-pharmaceutical intervention (NPI) can have an influence on COVID-19 mortality. At the time of variable inclusion, there were no published results for COVID-19 NPI yet. |
| Non-pharmaceutical intervention | npi_keystone_school_closure | Closure of schools and university | We expect non-pharmaceutical intervention (NPI) can have an influence on COVID-19 mortality. At the time of variable inclusion, there were no published results for COVID-19 NPI yet. |
| Non-pharmaceutical intervention | npi_keystone_shelter_in_place | An order indicating that people should shelter in | We expect non-pharmaceutical intervention (NPI) can have an influence on COVID-19 mortality. At |

| | | | |
|---|---|---|---|
| | | their homes except for essential reasons | the time of variable inclusion, there were no published results for COVID-19 NPI yet. |
| Non-pharmaceutical intervention | npi_keystone_social_distancing | Social distancing mandate of at least 6' between people | We expect non-pharmaceutical intervention (NPI) can have an influence on COVID-19 mortality. At the time of variable inclusion, there were no published results for COVID-19 NPI yet. |
| Socioeconomics | ses_hhincome | Median household income in the past 12 months | For most racial groups, increased income correlates with improved health.[29] |
| Socioeconomics | ses_pnohighschool | Percentage of no high school diploma by county | There is an established association of lower educational attainment and poorer health, including chronic illness and mortality,[30] along with the importance of high school education as a measure[31] of this association. |
| Socioeconomics | ses_ppoverty | Percentage of residents with income in the past 12 months below poverty level by county | For most racial groups, increased income correlates with improved health.[29] |
| Socioeconomics | ses_punemployed | Percentage of unemployed (not in labor force) by county | Unemployment is associated with poor health,[32] and also contributes to homelessness,[33] which is, in turn, a risk for COVID-19 infection.[34] |
| Social vulnerability | sv_groupquarterspop | Number of persons in institutionalized group quarters | This variable is used to construct the Social Vulnerability Index (SVI). Group living arrangements represent increased risk of SARS-CoV-2 transmission due to both difficulties in maintaining hygiene and social distancing in these settings and due to the risk that caregivers who visit multiple homes have an increased risk of both acquiring and spreading the virus.[35] |
| Social vulnerability | sv_p17below | Age 17 or younger | This variable is used to construct the Social Vulnerability Index (SVI). COVID-19 has a higher fatality rate for older populations, while sparing younger ages from more severe forms of the disease.[24] |

| | | | |
|---|---|---|---|
| Social vulnerability | sv_pcrowding | Percentage of occupied housing units with more people than rooms | This variable is used to construct the Social Vulnerability Index (SVI). This variable represents increased challenges to social distancing. For instance, an individual falling ill in a crowded apartment will have more difficulty in self-isolating than someone living in a spacious home. Individuals living in apartment complexes will have more difficulty in maintaining a 6-foot distance when outside than individuals with access to backyards.[36,37] |
| Social vulnerability | sv_pdisability | Older than age 5 with disability | This variable is used to construct the Social Vulnerability Index (SVI). This variable represents barriers to healthcare access and increased likelihood of greater health needs and worse outcomes from existing health conditions.[38] |
| Social vulnerability | sv_penglish | Percentage of population (age 5+) who speak English "less than well" | This variable is used to construct the Social Vulnerability Index (SVI). Populations with poor English skills are likely to have increased difficulty in accessing accurate health information and decreased visits to healthcare professionals.[39] |
| Social vulnerability | sv_pminority | Percentage minority (not non-Hispanic White) | This variable is used to construct the Social Vulnerability Index (SVI). The percentage of the county population who belongs to the racial minority is included for the same reason that different races/ethnicities were included as part of the demographic data above (Essentially, this is a grouping that includes all except non-Hispanic White. It can be understood as it might be better to use a composite variable to increase statistical power). Historically, more rural areas saw a lower burden of infectious disease because smaller populations meant diseases were less likely to be circulating,[23] suggesting that counties with smaller |

| | | | |
|---|---|---|---|
| | | | populations or larger land areas may be less impacted if those who are unwell come into contact with fewer people allowing the disease to burn out before cases and CFR climb. |
| Social vulnerability | sv_pmobilehome | Percentage of total housing units with mobile home | This variable is used to construct the Social Vulnerability Index (SVI). This variable represents increased challenges to social distancing. For instance, an individual falling ill in a crowded apartment will have more difficulty in self-isolating than someone living in a spacious home. Individuals living in apartment complexes will have more difficulty in maintaining a 6-foot distance when outside than individuals with access to backyards.[36,37] |
| Social vulnerability | sv_pmultiunit | Percentage of total housing units with 10 or more units | This variable is used to construct the Social Vulnerability Index (SVI). This variable represents increased challenges to social distancing. For instance, an individual falling ill in a crowded apartment will have more difficulty in self-isolating than someone living in a spacious home. Individuals living in apartment complexes will have more difficulty in maintaining a 6-foot distance when outside than individuals with access to backyards.[36,37] |
| Social vulnerability | sv_pnovehicle | Percentage of households with no vehicle available | This variable is used to construct the Social Vulnerability Index (SVI). Increased reliance on public transport will create more crowded transport and a higher risk of transmission.[40] |
| Social vulnerability | sv_singleparent | Single-parent household with children under 18 | This variable is used to construct the Social Vulnerability Index (SVI). These households are likely to experience increased difficulties finding childcare. The potential impact on absenteeism for |

| | | | |
|---|---|---|---|
| | | | healthcare workers could lead to higher mortality rates[41] if more of the workforce are single parents. |
| Demographics | demo_bridgedrace_p_ american_indians_alaskan | Percentage of (non Hispanic) American Indian or Alaska Native | In the US structural racism leads to racial/ethnic populations' lack of access to health care and receipt of low-quality health care, contributing to substantial health disparities,[42] which may, in turn, result in worse outcomes for COVID-19 patients. |
| Demographics | demo_bridgedrace_p_ asians_pacific | Percentage of (non-Hispanic) Asian or Pacific Islander. | In the US structural racism leads to racial/ethnic populations' lack of access to health care and receipt of low-quality health care, contributing to substantial health disparities,[42] which may, in turn, result in worse outcomes for COVID-19 patients. |
| Demographics | demo_bridgedrace_p_ blacks | Percentage of (non-Hispanic) Black or African American | In the US structural racism leads to racial/ethnic populations' lack of access to health care and receipt of low-quality health care, contributing to substantial health disparities,[42] which may, in turn, result in worse outcomes for COVID-19 patients. |
| Demographics | demo_bridgedrace_p_hisp | Percentage of Hispanic or Latino | In the US structural racism leads to racial/ethnic populations' lack of access to health care and receipt of low-quality health care, contributing to substantial health disparities,[42] which may, in turn, result in worse outcomes for COVID-19 patients. |
| Demographics | demo_bridgedrace_p_ whites | Percentage of (non-Hispanic) White | In the US structural racism leads to racial/ethnic populations' lack of access to health care and receipt of low-quality health care, contributing to substantial health disparities,[42] which may, in turn, result in worse outcomes for COVID-19 patients. |
| Demographics | demo_bridgedrace_total | Total population of each county (same as demo_population) | Historically, more rural areas saw a lower burden of infectious disease because smaller populations meant diseases were less likely to be circulating,[23] suggesting that counties with smaller populations or larger land areas may be less impacted if those who are unwell come into contact with fewer |

| | | | |
|---|---|---|---|
| | | | people allowing the disease to burn out before cases and CFR climb |
| Time | days_since_first_case | Number of days between first detected COVID19 case and final date of included case data, New York City data by county from NYC public health website, Kansas City counties were excluded | We expect this variable can have an influence on COVID-19 mortality. At the time of variable inclusion, there were no published results for this variable yet. |

**Appendix Exhibit A3: Variables and data sources**

| Model Inclusion Status | Variable Code | Level Data | Data Source | Year(s) Collected | Variable Unit Description |
|---|---|---|---|---|---|
| Included, linking variable | FIPS | County | US Census TIGER shapefile | 2018 | ID number |
| Excluded, highly correlated | como_allheartdis_hosp | County | CDC, Interactive Atlas of Heart Disease and Stroke | 2016-2018 | Incidence per 1000, 65+ |
| Excluded, highly correlated | como_allheartdis_mort | County | CDC, Interactive Atlas of Heart Disease and Stroke | 2014-2016 | Incidence per 100,000 |
| Included in final model | como_asthma | State | BRFSS | 2018 | Adjusted prevalence % |
| Excluded, non-significant in multivariate model | como_cancer5yr | County if available, o/w State | NIH, National Cancer Institute, State Cancer Profiles | 2012-2016 | 5-year incidence |
| Excluded, highly correlated | como_COPD | State | BRFSS | 2018 | Adjusted prevalence % |
| Excluded, highly correlated | como_cvd_hosp | County | CDC, Interactive Atlas of Heart Disease and Stroke | 2016-2018 | Incidence per 1000, ages 65+ |
| Excluded, highly correlated | como_cvd_mort | County | CDC, Interactive Atlas of Heart Disease and Stroke | 2016-2018 | Incidence per 100,000 |
| Excluded, non-significant in bivariate model | como_pdiabetes | County | CDC, US Diabetes Surveillance System | 2016 | Percentage (%) |
| Excluded, highly correlated | como_htn_hosp | County | CDC, Interactive Atlas of Heart Disease and Stroke | 2016-2018 | Incidence per 1000, 65+ |
| Excluded, non-significant in bivariate model | como_htn_mort | County | CDC, Interactive Atlas of Heart Disease and Stroke | 2016-2018 | Incidence per 1000 |
| Excluded, non-significant in multivariate model | como_ medicareheartdizprev | County | CDC, Interactive Atlas of Heart Disease and Stroke | 2018 | Prevalence per 1000, medicare beneficiaries |

| | | | | | |
|---|---|---|---|---|---|
| Excluded, highly correlated | como_pobesity | County | CDC, Interactive Atlas of Heart Disease and Stroke | 2016 | Percentage (%) |
| Excluded, non-significant in bivariate model | como_smoking | State | BRFSS | 2018 | Age adjusted prevalence |
| Excluded, highly correlated | como_stroke_hosp | County | CDC, Interactive Atlas of Heart Disease and Stroke | 2018 | Incidence per 1000 |
| Excluded, highly correlated | como_stroke_mort | County | CDC, Interactive Atlas of Heart Disease and Stroke | 2014-2016 | Incidence per 1000 |
| Excluded, non-significant in bivariate model | demo_landarea | County | US Census TIGER shapefile | 2018 | Area in square kilometer |
| Excluded, highly correlated | demo_p60more | County | Bridged race | 2010-2018 average | Percentage (%) |
| Included in final model | demo_p65more | County | Bridged race | 2010-2018 average | Percentage (%) |
| Excluded, highly correlated | demo_p45_64 | County | Bridged race | 2010-2018 average | Percentage (%) |
| Excluded, highly correlated | demo_popdensity | County | Bridged race | 2018 | Person per square kilometer |
| Excluded, highly correlated | demo_population | County | Bridged race | 2018 | Count |
| Included in final model | hc_hospitals | County | CDC, Interactive Atlas of Heart Disease and Stroke and demo_population variable | 2018 | Total number of hospitals in county |
| Included in final model | hc_hospitals_per10000 | County | CDC, Interactive Atlas of Heart Disease and Stroke and demo_population variable | 2018 | Number of hospitals per 10,000 people in county |
| Excluded, highly correlated | hc_icubeds_per1000 | County | Kaiser Health News analysis of hospital cost reports filed to the Centers for Medicare | 2018/2019 | Number per 1000 persons in the county |

| | | | | | |
|---|---|---|---|---|---|
| | | | & Medicaid Services American Community Survey, (5-year estimate) | | |
| Excluded, highly correlated | hc_icubeds | County | Kaiser Health News analysis of hospital cost reports filed to the Centers for Medicare & Medicaid Services American Community Survey, (5-year estimate) | 2018/2019 | Number of ICU beds in county |
| Excluded, non-significant in multivariate model | hc_icubeds_per60more | County | Kaiser Health News analysis of hospital cost reports filed to the Centers for Medicare & Medicaid Services | 2018/2019 | Number per 1000 persons aged 60+ |
| Excluded, highly correlated | hc_icubeds_per65more | County | Kaiser Health News analysis of hospital cost reports filed to the Centers for Medicare & Medicaid Services | 2018/2019 | Number per 1000 persons aged 65+ |
| Excluded, highly correlated | hc_medicaid | County | CDC, Interactive Atlas of Heart Disease and Stroke | 2018 | Percentage (%) |
| Included in final model | hc_Pnotinsured_acs | County | US Census American Community Survey 5-Year Data | 2018 | Percentage (%) |
| Excluded, non-significant in bivariate model | hc_primarycare | County | Health Resources and Services Administration, (Area Health Resources File) | 2016 | Count |
| Excluded, highly correlated | hc_primarycare_per1000 | County | Health Resources and Services Administration, (Area Health Resources File) | 2016 | Adjusted incidence rate per 1000 |
| Excluded, highly correlated | npi_keystone_closing_of_public_venues | County | KeyStone Coronavirus City and County Non-Pharmaceutical Intervention Rollout Date Dataset | 2020 | Date |

| | | | | | |
|---|---|---|---|---|---|
| Excluded, highly correlated | npi_keystone_gathering_size_10_0 | County | KeyStone Coronavirus City and County Non-Pharmaceutical Intervention Rollout Date Dataset | 2020 | Date |
| Excluded, highly correlated | npi_keystone_gathering_size_100_to_26 | County | KeyStone Coronavirus City and County Non-Pharmaceutical Intervention Rollout Date Dataset | 2020 | Date |
| Excluded, highly correlated | npi_keystone_gathering_size_25_to_11 | County | KeyStone Coronavirus City and County Non-Pharmaceutical Intervention Rollout Date Dataset | 2020 | Date |
| Excluded, highly correlated | npi_keystone_gathering_size_500_to_101 | County | KeyStone Coronavirus City and County Non-Pharmaceutical Intervention Rollout Date Dataset | 2020 | Date |
| Excluded, highly correlated | npi_keystone_lockdown | County | KeyStone Coronavirus City and County Non-Pharmaceutical Intervention Rollout Date Dataset | 2020 | Date |
| Excluded, highly correlated | npi_keystone_non_essential_services_closure | County | KeyStone Coronavirus City and County Non-Pharmaceutical Intervention Rollout Date Dataset | 2020 | Date |
| Excluded, highly correlated | npi_keystone_Other | County | KeyStone Coronavirus City and County Non-Pharmaceutical Intervention Rollout Date Dataset | 2020 | Date |
| Included in final model | npi_keystone_religious_gatherings_banned | County | KeyStone Coronavirus City and County Non-Pharmaceutical Intervention Rollout Date Dataset | 2020 | Date |

| | | | | | |
|---|---|---|---|---|---|
| Excluded, highly correlated | npi_keystone_school_closure | County | KeyStone Coronavirus City and County Non-Pharmaceutical Intervention Rollout Date Dataset | 2020 | Date |
| Excluded, highly correlated | npi_keystone_shelter_in_place | County | KeyStone KeyStone Coronavirus City and County Non-Pharmaceutical Intervention Rollout Date Dataset | 2020 | Date |
| Excluded, non-significant in multivariate model | npi_keystone_social_distancing | County | KeyStone Coronavirus City and County Non-Pharmaceutical Intervention Rollout Date Dataset | 2020 | Date |
| Excluded, highly correlated | ses_hhincome | County | US Census American Community Survey 5-Year Data | 2018 | Median income in US Dollars |
| Excluded, highly correlated | ses_pnohighschool | County | US Census American Community Survey 5-Year Data | 2018 | Percentage (%) |
| Excluded, highly correlated | ses_ppoverty | County | US Census American Community Survey 5-Year Data | 2018 | Percentage (%) |
| Excluded, non-significant in multivariate model | ses_punemployed | County | US Census American Community Survey 5-Year Data | 2018 | Percentage (%) |
| Excluded, highly correlated | sv_groupquarterspop | County | US Census American Community Survey 5-Year Data | 2018 | Count |
| Excluded, highly correlated | sv_p17below | County | US Census American Community Survey 5-Year Data | 2018 | Percentage (%) |

| | | | | | |
|---|---|---|---|---|---|
| Excluded, non-significant in multivariate model | sv_pcrowding | County | US Census American Community Survey 5-Year Data | 2018 | Percentage (%) |
| Excluded, highly correlated | sv_pdisability | County | US Census American Community Survey 5-Year Data | 2018 | Percentage (%) |
| Excluded, highly correlated | sv_penglish | County | US Census American Community Survey 5-Year Data | 2018 | Percentage (%) |
| Excluded, highly correlated | sv_pminority | County | CDC SVI | 2018 | Percentage (%) |
| Included in final model | sv_pmobilehome | County | US Census American Community Survey 5-Year Data | 2018 | Percentage (%) |
| Excluded, highly correlated | sv_pmultiunit | County | US Census American Community Survey 5-Year Data | 2018 | Percentage (%) |
| Excluded, non-significant in multivariate model | sv_pnovehicle | County | US Census American Community Survey 5-Year Data | 2018 | Percentage (%) |
| Excluded, highly correlated | sv_singleparent | County | US Census American Community Survey 5-Year Data | 2018 | Percentage (%) |
| Excluded, non-significant in bivariate model | demo_bridgedrace_p_american_indians_alaskan | County | CDC, National Center for Health Statistics | 2010-2018 | Percentage (%) |
| Excluded, non-significant in bivariate model | demo_bridgedrace_p_asians_pacific | County | CDC, National Center for Health Statistics | 2010-2018 | Percentage (%) |
| Included in final model | demo_bridgedrace_p_blacks | County | CDC, National Center for Health Statistics | 2010-2018 | Percentage (%) |
| Excluded, highly correlated | demo_bridgedrace_p_hisp | County | CDC, National Center for Health Statistics | 2010-2018 | Percentage (%) |

| | | | | | |
|---|---|---|---|---|---|
| Excluded, highly correlated | demo_bridgedrace_p_whites | County | CDC, National Center for Health Statistics | 2010-2018 | Percentage (%) |
| Excluded, highly correlated | demo_bridgedrace_total | County | CDC, National Center for Health Statistics | 2010-2018 | Percentage (%) |
| Excluded, highly correlated | days_since_first_case | County | NYT COVID-19 Dataset | Jan 21-Jun 12, 2020 | Days |

**Appendix Exhibit A4: Lag adjusted case-fatality rate (laCFR) calculation**

During the first wave of the pandemic, SARS-CoV-2 was non-endemic, leading the case-fatality rate (CFR) to fluctuate over time. This is due to a lag when counting the number of deaths compared to cases and hospitalizations, leading to an underestimation of the CFR. The CFR continues to fluctuate rapidly early in an epidemic when each additional case or death has an excessive impact on calculating CFR. It is important to not only account for the lag between cases and deaths (i.e., lag-adjusted CFR), but also to ensure that the CFR is no longer fluctuating.

To do this, we use a method developed by Nishiura et al. and expanded upon by Russell et al., where case and death incidence data are used to estimate the number of cases with known outcomes, i.e. cases where the resolution, death or recovery, is known to have occurred:[43,44]

$$u_t = \frac{\sum_{i=0}^{t} \sum_{j=0}^{\infty} c_{i-j} f_i}{\sum_{i=0}^{t} c_j}$$

where $c_t$ is the daily case incidence at time $t$, (with time measured in calendar days), $f_t$ is the proportion of cases with delay $t$ between onset or hospitalization and death; $u_t$ represents the underestimation of the known outcomes and is used to scale the value of the cumulative number of cases in the denominator in the calculation of the laCFR. Russell et al. used the estimated distribution in Linton et al., based on data from China up until the end of January 2020. For this study, we instead used United States centric data from Lewnard et al., which estimates the distribution of time from hospitalization to death based on data from Washington and California.[45]

Lewnard et al., fits the distribution conditionally on age resulting in a Weibull distribution for each age group.[45] The overall distribution was obtained empirically by weighting the densities at time $t$ across all age groups. Because of this, the overall distribution doesn't have its own shape/scale parameters. However, we were able to estimate what these parameters would be by fitting a Weibull distribution that captures the 2.5, 25, 50, 75, and 97.5 percentiles, as well as the average.

Use of the laCFR assumes the measure has stabilized.[43] Counties where the laCFR is still rapidly changing cannot be used in the study as these are not unbiased estimates of the true CFR. laCFRs were calculated incrementally for each day and assessed whether they changed on average less than 1% a week for the last two weeks of available data. The final calculation based on all data available was used as the laCFR in our model.

**Appendix Exhibit A5: Descriptive statistics for county variables retained for analysis (median and range for continuous variables)**

| Variable Code | Training Set (n=1186) | | Testing Set (n=593) | | Excluded (n=1364) | |
|---|---|---|---|---|---|---|
| como_asthma (%) | 9.4 | (7.4–12.8) | 9.2 | (7.4–12.8) | 9.2 | (7.4–12.8) |
| como_cancer5yr (cases/5yr) | 463.3 | (272.1–1135) | 457.8 | (241–592.1) | 451.0 | (130.1–677.2) |
| como_pdiabetes (%) | 10.1 | (1.5–33.0) | 10.0 | (1.7–24.6) | 9.5 | (1.8–32.3) |
| como_htn_mort (deaths/100,000) | 120.2 | (20.4–400.6) | 120.2 | (18.7–442.2) | 129.7 | (26.5–592.1) |
| como_medicareheartdizprev (%) | 36.0 | (22.1–55.2) | 35.7 | (19.5–49.3) | 35.3 | (18.0–53.5) |
| como_smoking (%) | 17.3 | (9.0–26.8) | 17.3 | (9.0–26.8) | 17.7 | (9.0–26.8) |
| demo_landarea ($m^2$) | 1535.8 | (6.5–64008) | 1505.1 | (38.8–51954) | 1743.8 | (5.3–377034) |
| demo_p65more (%) | 15.8 | (4.2–51.5) | 15.6 | (6.5–27.6) | 19.1 | (5.1–36.1) |
| demo_bridgedrace_p_american_indians_alaskan (%) | 0.3 | (0.0–73.4) | 0.3 | (0.1–92.2) | 0.5 | (0.0–93.9) |
| demo_bridgedrace_p_asians_pacific (%) | 0.9 | (0.1–65.5) | 1.0 | (0.1–30.4) | 0.5 | (0.0–59.1) |
| demo_bridgedrace_p_blacks (%) | 4.6 | (0.1–82.6) | 5.5 | (0.2–78.5) | 1.1 | (0.0–85.7) |
| hc_hospitals (hospitals) | 1 | (0–32) | 1 | (0–79) | 1 | (0–8) |
| hc_hospitals_per10000 (hospitals/10,000) | 0.2 | (0.0–3.8) | 0.2 | (0.0–4.7) | 0.4 | (0.0–8.5) |
| hc_icubeds_per60more (beds/>60yr resident) | 0.6 | (0.0–8.2) | 0.7 | (0.0–7.0) | 0.0 | (0.0–101.1) |
| hc_pnotinsured_acs (%) | 9.0 | (1.8–39.2) | 9.4 | (2.0–35.6) | 9.3 | (1.7–45.6) |
| hc_primarycare (physicians) | 1.9 | (0.2–46.6) | 1.8 | (0.4–17.9) | 2.3 | (0.2–19.9) |
| npi_keystone_religious_gatherings_banned (% counties that ban religious gatherings) | 52.4 | | 54.8 | | 47.3 | |
| npi_keystone_social_distancing (days since 1$^{st}$ county case) | 3 | (-46–76) | 3 | (-30–82) | -8 | (-84–46) |
| ses_punemployed (%) | 3.3 | (0.5–9.5) | 3.3 | (0.5–13.6) | 2.9 | (0.0–16.5) |
| sv_pcrowding (%) | 0.8 | (0.0–10.1) | 0.9 | (0.0–15.4) | 0.9 | (0.0–35.5) |
| sv_pmobilehome (%) | 9.5 | (0.0–54.8) | 8.7 | (0.0–51.2) | 12.2 | (0.0–59.3) |
| sv_pnovehicle (%) | 5.8 | (1.4–77.0) | 5.9 | (1.0–32.2) | 5.4 | (0.0–87.8) |

**Appendix Exhibit A6: Regression results**

| Variable | Exp (Coeff.) | 95% CI | | Std. Error | Wald | *p*-value | VIF |
|---|---|---|---|---|---|---|---|
| Intercept | 0.0111 | (0.0062, | 0.0200) | 0.2961 | -15.2016 | <0.0001 | |
| Hospitals per 10,000 | 0.6773 | (0.5549, | 0.8248) | 0.1013 | -3.8485 | 0.0001 | 1.0636 |
| Religious Gatherings Ban | 0.8752 | (0.7894, | 0.9702) | 0.0526 | -2.5315 | 0.0114 | 1.0592 |
| Pop. Not Insured (%) | 0.9855 | (0.9715, | 0.9998) | 0.0075 | -1.9444 | 0.0519 | 1.6001 |
| Mobile Home Pop. (%) | 0.9921 | (0.9854, | 0.9989) | 0.0035 | -2.2539 | 0.0242 | 1.7227 |
| Asthma Pop. (%) | 1.0951 | (1.0400, | 1.1533) | 0.0264 | 3.4378 | 0.0006 | 1.1489 |
| Pop. >= 65 Yrs. (%) | 1.0453 | (1.0308, | 1.0605) | 0.0069 | 6.4394 | <0.0001 | 1.1405 |
| Total Hospitals in County | 1.0316 | (1.0100, | 1.0522) | 0.0099 | 3.1329 | 0.0017 | 1.1663 |
| Black Pop. (%) | 1.0097 | (1.0063, | 1.0133) | 0.0018 | 5.5012 | <0.0001 | 1.2210 |

County-level predictors of COVID-19 cCFR in the United States, $R^2 = 0.8620$.

**Appendix Exhibit A7: Model fit**

We compared the mean and variance seen within our model predictions to the theoretical mean and variance expected in a Poisson and negative binomial model. After grouping the fitted predictions into 20 quantiles and calculating their means and variances, we saw the negative binomial model captures our data variance well.[46] Loess smooth was used for the empirical mean (Exhibit 2 (A)). As an additional check, we calculated the ratio of Pearson residuals to degrees of freedom, which was 1.04, indicating we accounted for most of the over-dispersion in cCFR using the negative binomial model. This was confirmed with a half-normal plot (Exhibit 2 (B)). The simulated envelope for the deviance residuals in the half-normal plot serves as a guide of what to expect under a well-fitted model, with most of our model's deviance residuals lying within.[47] The Cox and Snell Pseudo R2 for our model was 0.86, which accounts for the majority of the variance present in our outcome variable. All variables had a variance inflation factor of less than 2, indicating collinearity was not an issue with our variables (Appendix A4).

Similar to ROC, a gain curve plot measures how well the model score sorts the data compared to the true outcome value.[48] When the predictions sort in exactly the same order, the relative Gini coefficient is 1. When the model sorts poorly, the relative Gini coefficient is close to zero, or even negative. The relative Gini scores were high for both our training set and testing set. (0.9840 and 0.9829, respectively, Exhibit 2 (D)). We also checked the coverage, which is the probability that our model outcomes are found within our prediction interval. To estimate our predictive coverage (empirical coverage), we simulated a prediction interval. The coverage was 0.9730 for the training data and 0.9713 for testing data (Exhibit 2 (C)).
